# On the COVID-19 Pandemic in Indian State of Maharashtra: Forecasting & Effect of different parameters

**DOI:** 10.1101/2020.05.23.20111179

**Authors:** Arvind S. Avhad, Prasad P. Sutar, Onkar T. Mohite, Vaibhav S. Pawar

## Abstract

This work details the outbreak and factors affecting the spread of novel coronavirus (COVID-19) in the Indian state of Maharashtra, which is considered as one of the most massive and deadly pandemic outbreaks. Observational data collected between 14 March 2020 and 4 May 2020 is statistically analyzed to determine the nonlinear behavior of the epidemic. It is followed by validating predicted results with real-time data. Proposed model is further used to obtain statistical summaries in which Grubbs tests for outlier detection have justified high values of evaluation metrics. Outliers are found to be pilot elements in an outbreak under considered region. Statistically, a significant correlation has been observed between dependent and explanatory variables. Transmission pattern of this virus is very much different from the SARS-CoV-1 virus. Key findings of this work will be predominant in maintaining environment conditions at healthcare facilities to reduce transmission rates at these most vulnerable places.

## 1. Introduction

The first case of COVID-19, a new coronavirus disease in the Indian state of Maharashtra was reported on 9 March 2020 with a fatality rate of 4.00 %, significantly higher than the rest of the country with a confirmed total of 14,541 cases, including 2,465 recoveries and 582 deaths as of 4 May 2020. Researchers have begun concentrating on the outbreak of novel viruses using Data Science tools to forecast, predict, and learn about this novel virus. Besides, some used statistical techniques to forecast and study factors governing the spread of the virus. (Dangi, et al., March 2020) [1]Predicted the potential spread of COVID-19 through an analytical study based on temperature, population, and longitudinal analysis for selected foreign cities where the COVID-19 outbreak is pandemic. Their work provides a strong positive correlation between selected international cities and Indian national cities. (Fanelli, et al., 2020) [2] Analyzed and foretold the range of COVID-19 in China, Italy, and France using nonlinear fitting of the population model. The data analyzed in this study stretches between 22 January 2020, and 15 March 2020.

(Ghazaly, et al., April 2020) [3] Predicted outbreak of coronavirus through time series using non-linear regressive network (NAR) using artificial intelligence and deep learning. The forecast is done for nine countries namely; Egypt, Saudi Arabia, Jordan, United States Of America, Spain, Italy, France, Iran, Russian Federation. (Ivanov, 2020) [4] presents the results of simulation study that opens some new research tensions on the impact of COVID-19 on the global SCs. (Jianxi Luo, April 2020) [5] predicted not only the subsequent developments of the COVID-19 outbreak but also end dates in different countries. In their study, analysis is done through regression, which runs for individual countries with the newest data. The regressed model is used to estimate the full pandemic cycle and plot the life cycle curve.

(Petropoulos, et al., 2020) [6] Introduces an objective approach to predict the continuation of COVID-19. This work describes the timeline of live forecasting exercise with massive potential applications for planning and decision making. (Remuzzi, April 2020) [7] Analyzes COVID-19 outbreak in Italy which might help political leaders and health authorities to allocate enough resources, including personnel, beds, and intensive care facilities, to manage the situation. (Wei-Jie-guan, April 2020) [8] Proposed the clinical characteristics of coronavirus disease 2019 in china through statistical analysis. Continuous variables were expressed as medians and ranges using R software. (Xu X, et al., 2020)[9] Points to the important discovery that RBD domain of the Wuhan CoV S-protein supports strong interaction with human ACE2 molecules despite its sequence diversity with SARS-CoV S-protein.

In this paper, the nonlinear regression analysis is applied to analyze the coronavirus outbreak problem in the state of Maharashtra with data available from the government’s official corona update portal and related sources. Nonlinear regression analysis models the observational data by functions, which are the nonlinear combinations of the model parameters considered in outbreak analysis. The output of this analysis depends on multiple independent variables. Successive approximations are used to obtain the best fit of observational data. The model analyzes different variables associated with the outbreak of the novel coronavirus, which represents its regression and predicts the impact on the spread of the disease. Objectives of the paper are: (a) Forecasting the outbreak in Maharashtra till the end of July 15, 2020; (b) Investigating the sensitivity of the confirmed cases to different parameters, viz. temperature, humidity, tests etc. and (c) Using this analysis to prevent an outbreak and gaining some insight for vaccine development.

## 2. Methods

In this study, the evaluation model for the outbreak of COVID-19 coronavirus is developed to study the impact of various factors associated with the outbreak and use them to forecast the outbreak in the state of Maharashtra, as shown in Fig. 1. The proposed region of the state has been divided into 36 sub-regions categorized as Red, Orange, and Green zones as per Government of India directives, as shown in Fig. 2.

**Fig. 1.**
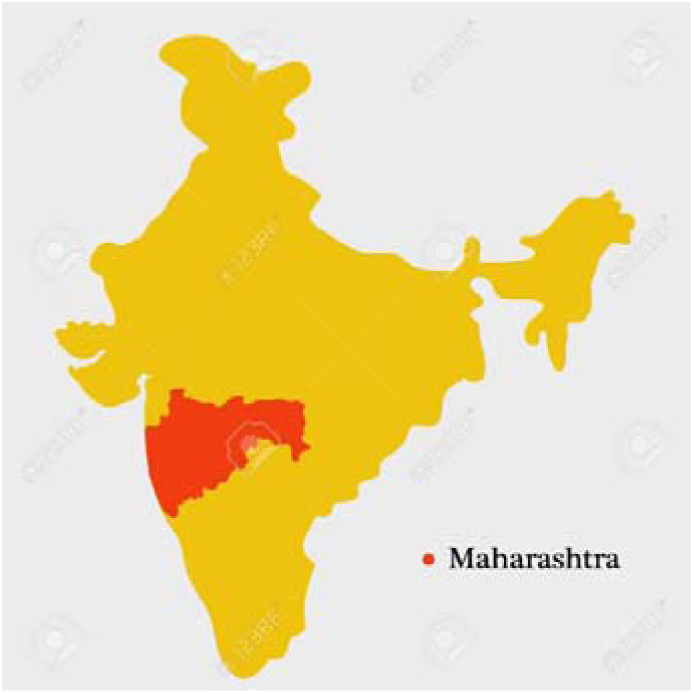
Selected region.

**Fig. 2.**
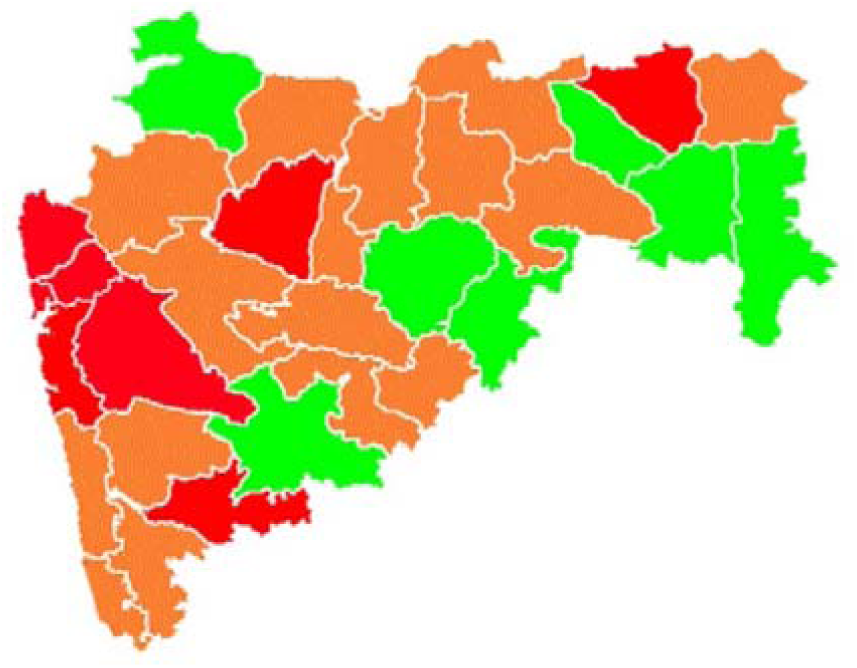
Zones.

Data collection is done from the government of Maharashtra COVID-19 portal from 14 March 2020 to 4 May 2020. Other parameters related to factors affecting outbreaks are taken from the government’s official sources. The total sample collected for work is divided into two parts, one for the evaluation as well as prediction of outbreak characteristic and other for the forecasting. Data are summarized in Table 1.

**Table 1.** Sample of data collection [10] [11].

| DATE | CONFIRMED | ACTIVE | RECOVERED | DECEASED |
| --- | --- | --- | --- | --- |
| 14-Mar | 14 | 14 | 0 | 0 |
| 15-Mar | 32 | 32 | 0 | 0 |
| 01-Apr | 335 | 283 | 39 | 13 |
| 27-Apr | 8590 | 6940 | 1282 | 484 |
| 4-May | 14541 | 11494 | 2465 | 582 |

XLSTAT statistical analysis tool is used to build, evaluate, and prepare the predictions of the model. SSE, MSE, and AIC are chosen as evaluation metrics for regression models as R-squared alone cannot be trusted for estimation of a good fit of data since nonlinear functions are highly flexible to fit into complex curves. The model that fits the data with the best values for the chosen standard of measurements is considered as an optimal model. Of all the variables initially considered, to get the best set of variables, the forward stepwise method is selected. This method enables us to overcome the time-consuming selection process and the requirement of extensive computation capabilities. As the factors governing the corona outbreak are more, the regression model becomes multivariate.

The algorithm used in present work works on successive iteration with an increasing number of independent variables and creating models for each combination. The model that best fits with evaluation metrics is selected for statistical analysis.

## 3. Results and discussion

Observational data is evaluated as a simple linear regression model, multivariate linear regression model, and nonlinear regression model with considered explanatory variables and confirmed cases as the dependent variables. The best suitable explanatory variables subset is the temperature (month of April average), humidity (month of April average), and the approximate number of tests carried out (1297/ million population). Following algorithm as shown in the Fig. 3 and easy-fit test between linear regression and random forest test, the possibility of analysis to be linear regression is ruled out based upon results shown in Table 2. For this purpose, data selected were randomly separated into two samples. 80% of the observations were used to train the model and 20% for validation.

**Fig. 3.**
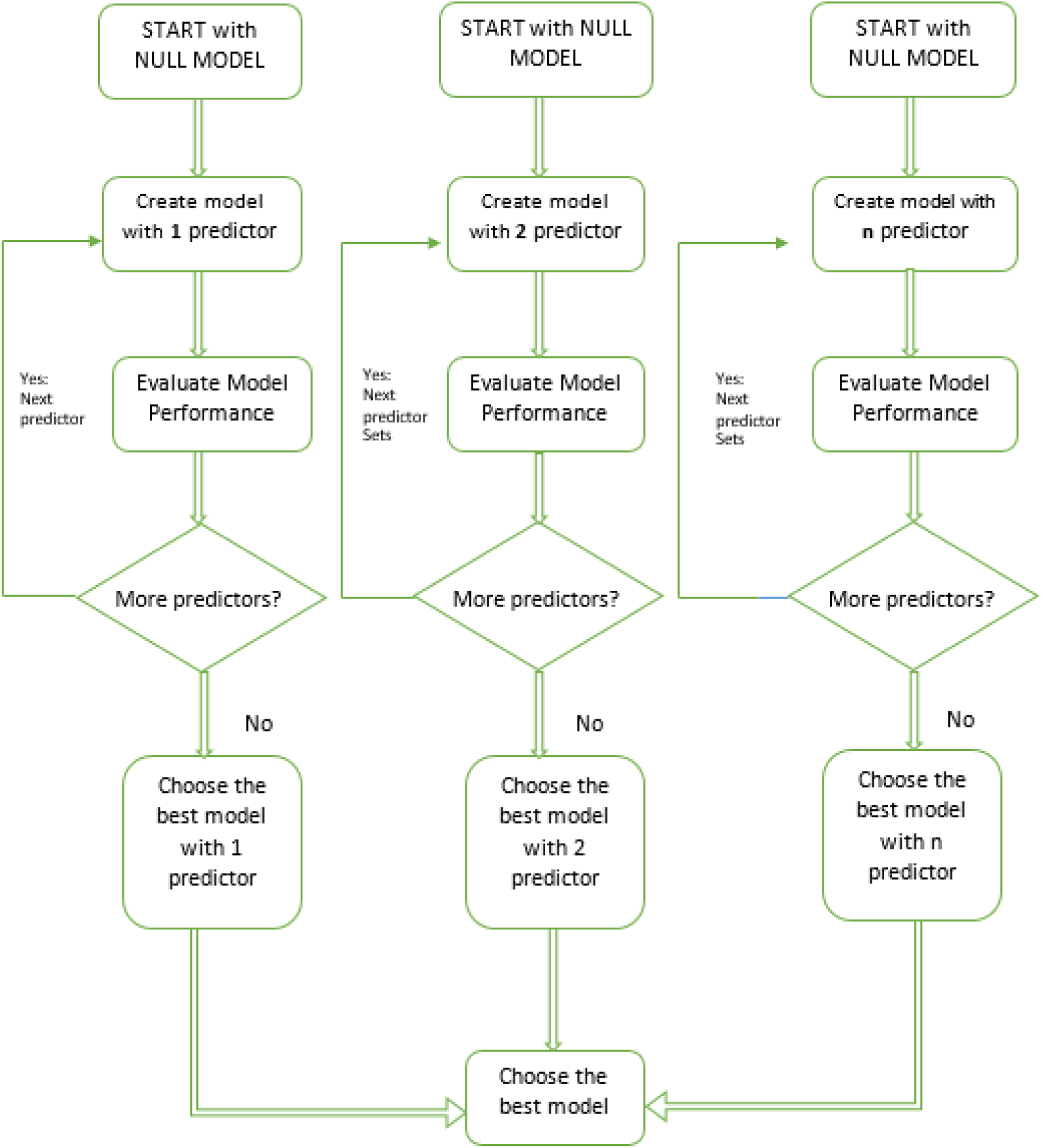
Algorithm for selection of model.

**Table 2.** AI Easy-fit Tool (Summary table:)

|  | Linear regression | Random forests |
| --- | --- | --- |
| Mean squared error (Validation) | 337929.316 | 5433.181 |
| *The best model, according to the statistic Mean squared error computed on the validation sample, is coloured in green in the table above.* |  |  |

Further proceeding with nonlinear regression, the selected model was evaluated for the polynomial function of Second-order, third-order, and so on with goodness of fit, as shown in Fig. 4.

**Fig. 4.**
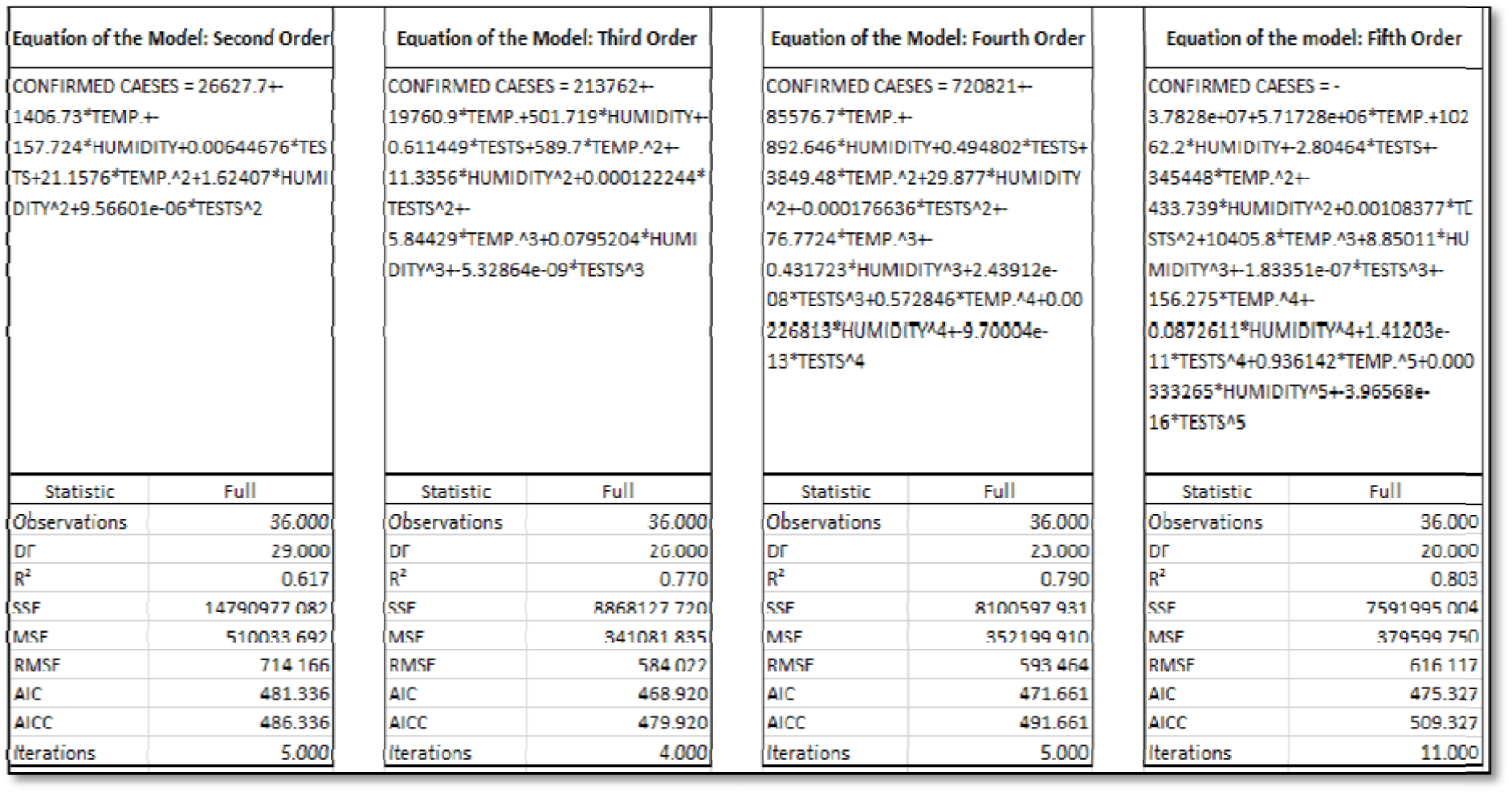
Goodness of fit.

It is observed that evaluation metrics, such as R^2^ value is lower, and errors, as well as AIC, are smaller at second-order polynomial function. In the third order, there is a sudden rise in R^2^ value and a decrease in other metrics. Further proceeding with higher orders, it is evident that increases in the R^2^ value is not very high and errors, as well as AIC values, start increasing. This implies that the third-order polynomial regression is best among all polynomial models evaluated.

Data for confirmed COVID-19 positive cases reported is collected from 14 March 2020, to 4 May 2020. Based on goodness of fit analysis, predictions of confirmed cases with third-order polynomial are made as shown in Fig. 5-8.

**Fig. 5.**
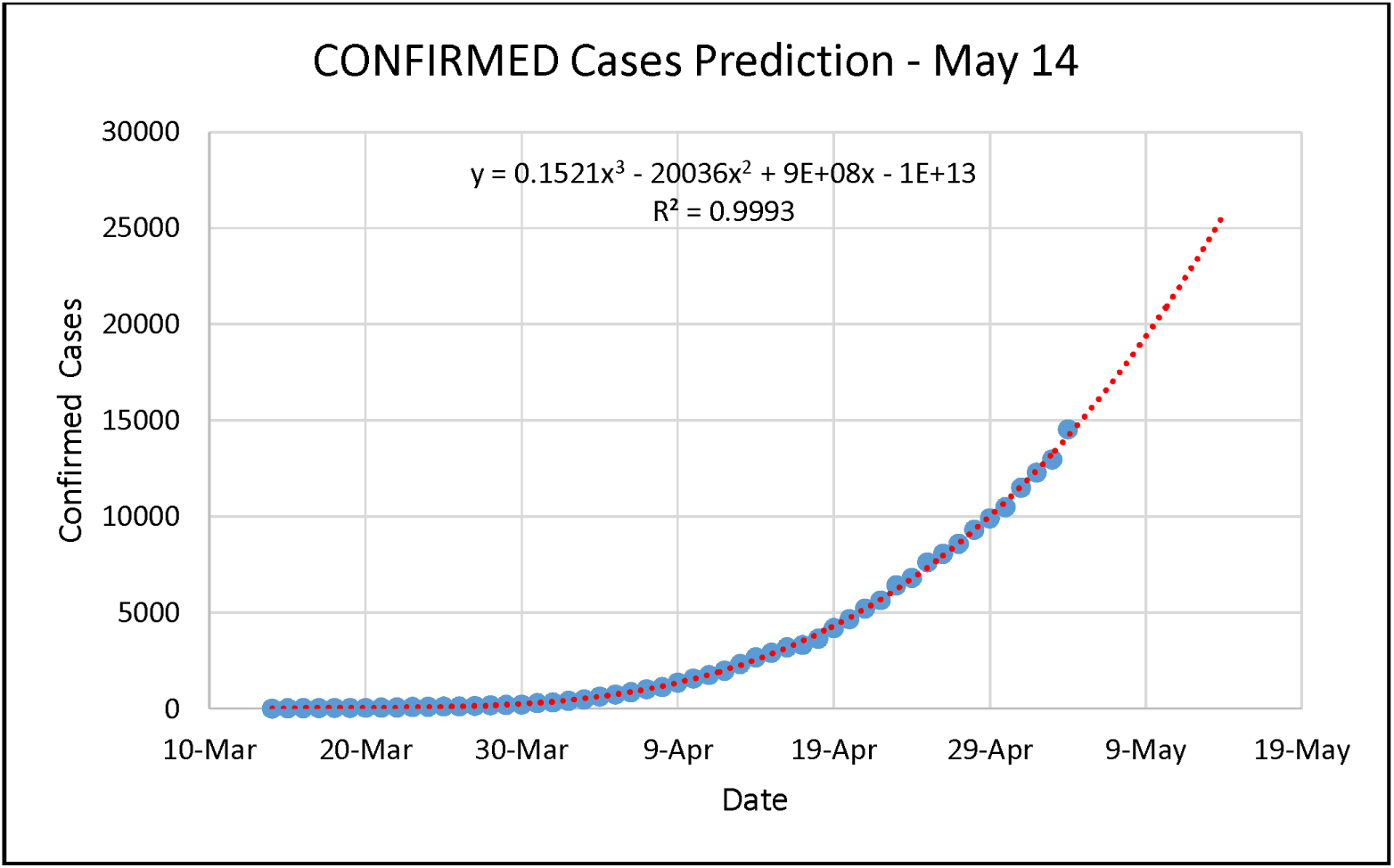
Prediction 14 May.

**Fig. 6.**
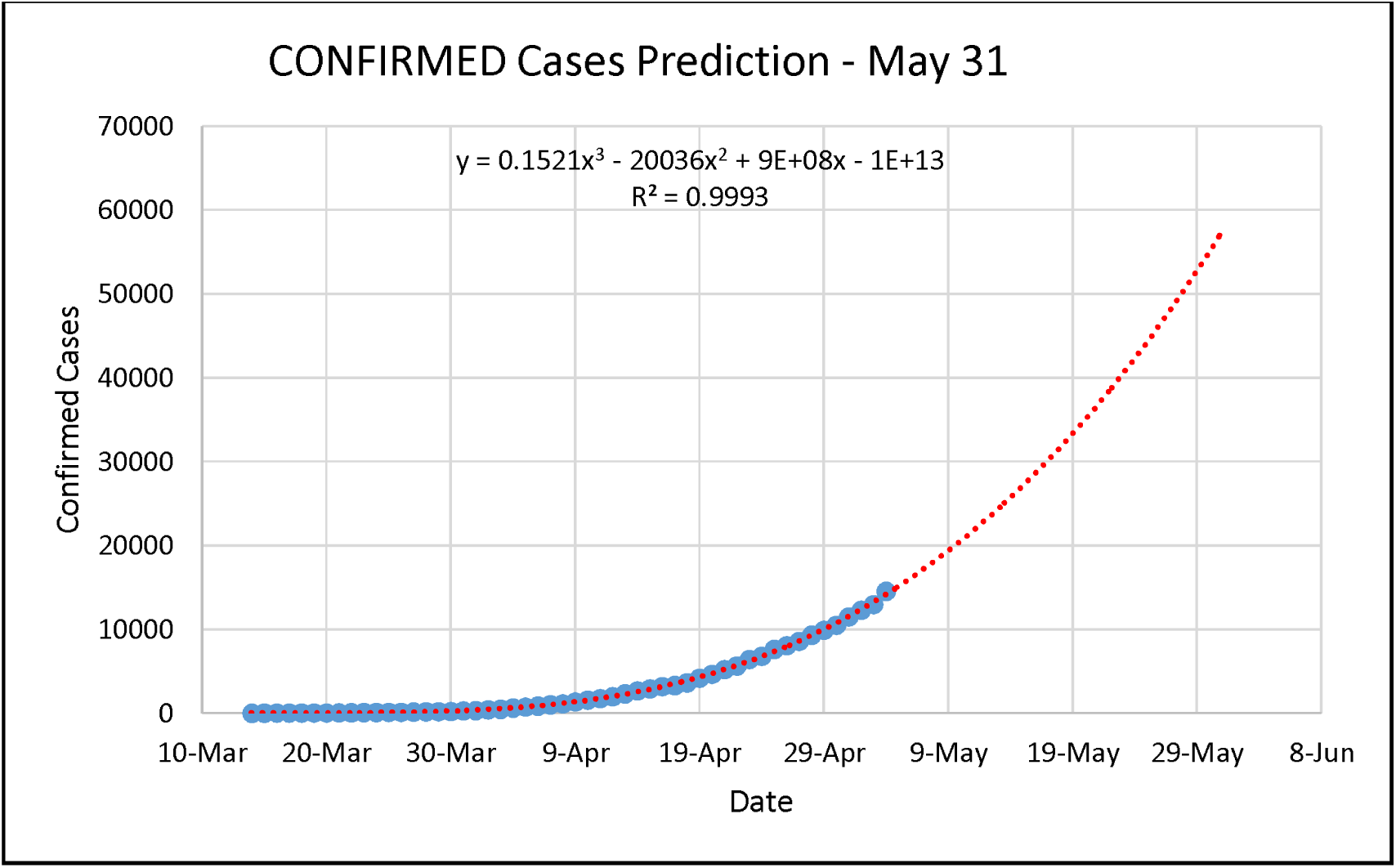
Prediction 31 May.

**Fig. 7.**
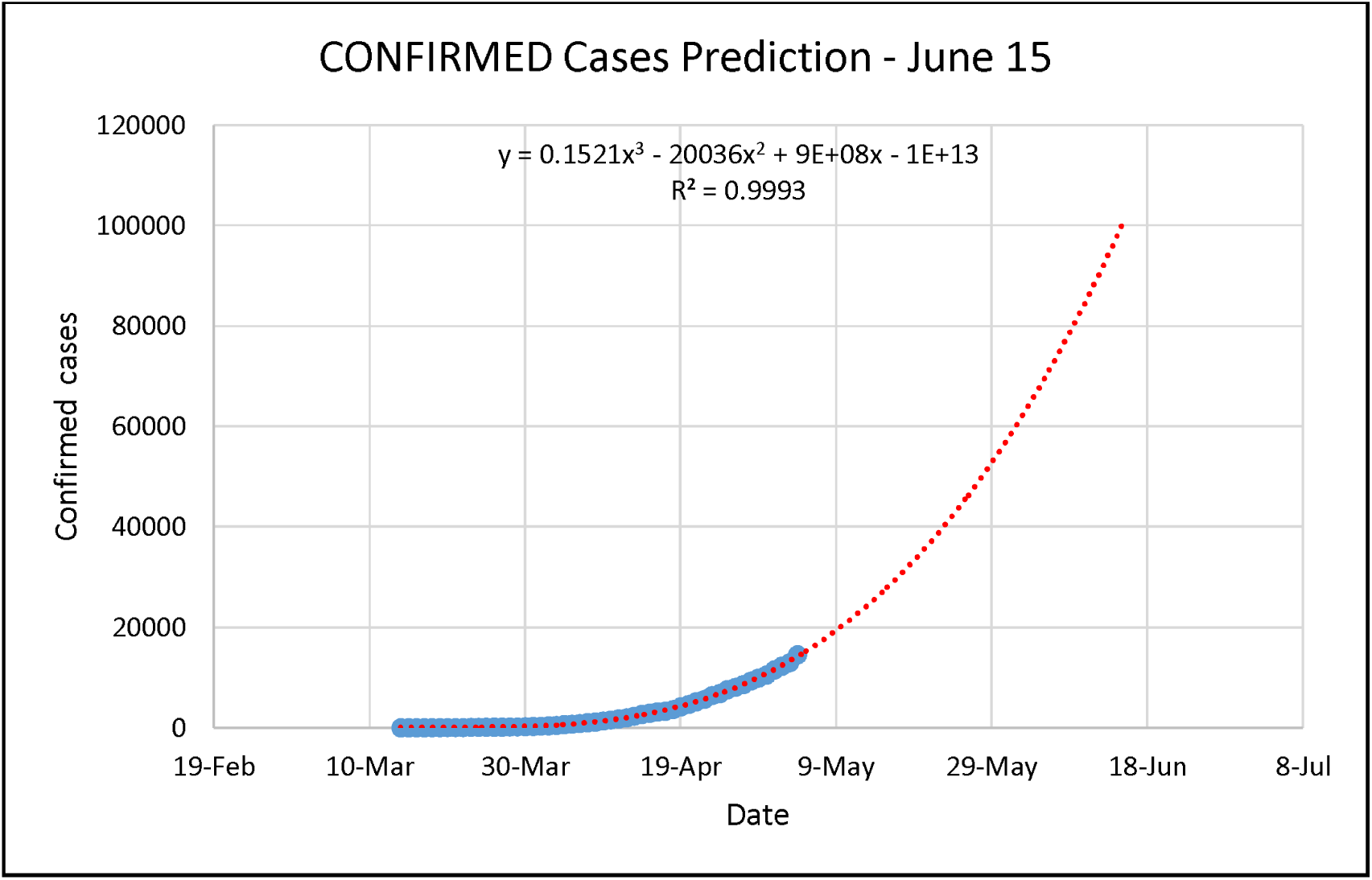
Prediction 15 June.

**Fig. 8.**
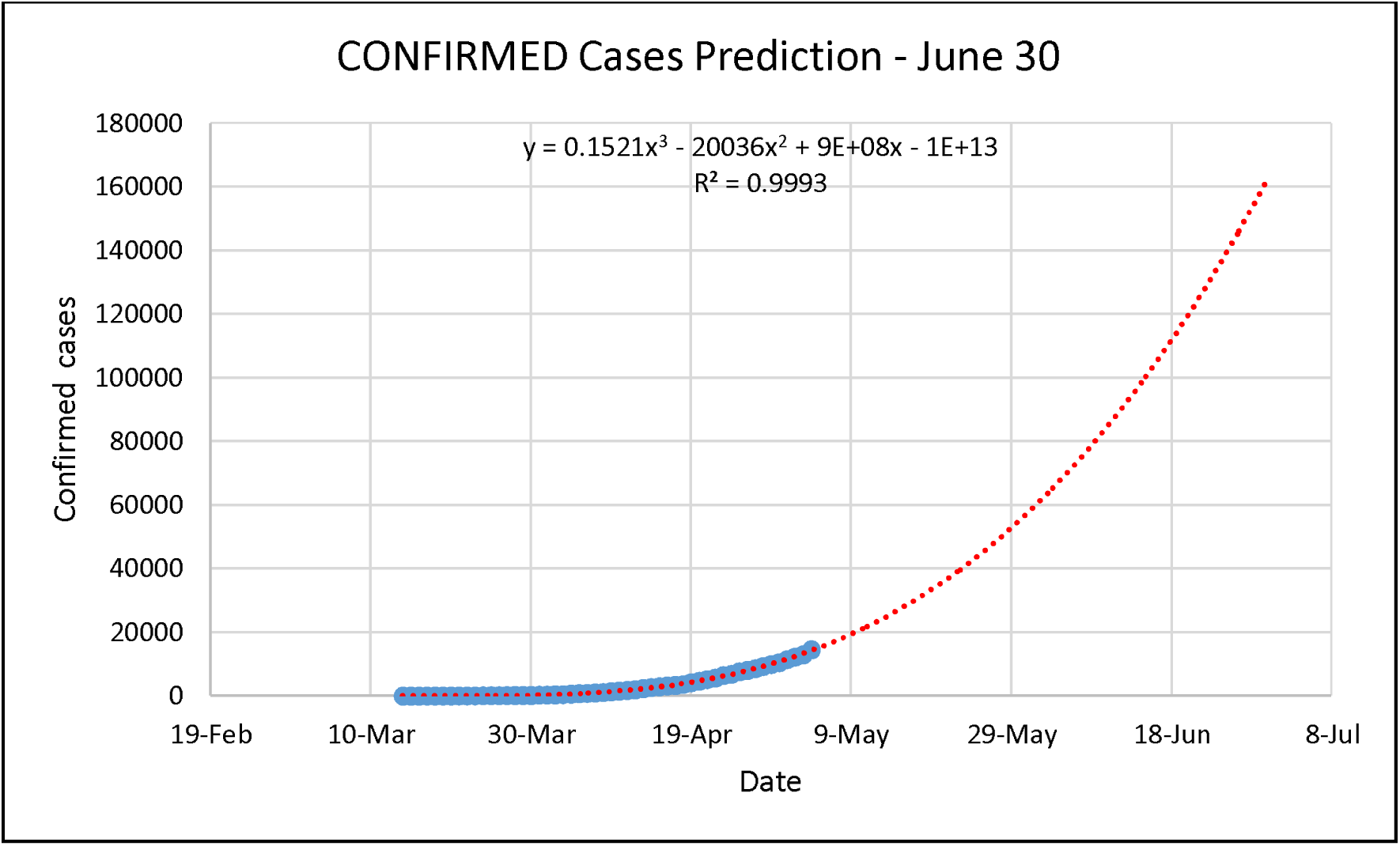
Prediction 30 June.

From Fig.5 the predicted number of confirmed cases for data in Maharashtra on May 14^th^ is around 26000. The actual number of confirmed cases reported on May 14^th^ is 27524 [11] thus predicted, the number of confirmed cases significantly corresponds with the exact amount of cases reported when the forecast is made with a third-order polynomial. Hence the choice of third-order polynomial regression through statistical modelling is validated against real data.

Predictions, as shown in Fig.5 and Fig. 6 are based on data collected between, March 14^th^ 2020 and May 4^th^ 2020 (nationwide lockdown period).

**[Note: May 5^th^ onwards, certain relaxations in lockdown are permitted. Thus, with partial lockdown, when the data is considered till May 14^th^ from 14^th^ March, predicted number of confirmed cases are 64000 at the end of May 31^st^ 2020, excellent agreement with the data available on government portal; 116000 by the end of June 15, 185000 (till end of June 30), 285000 by end of July 15 etc.]**

**Fig. 9.**
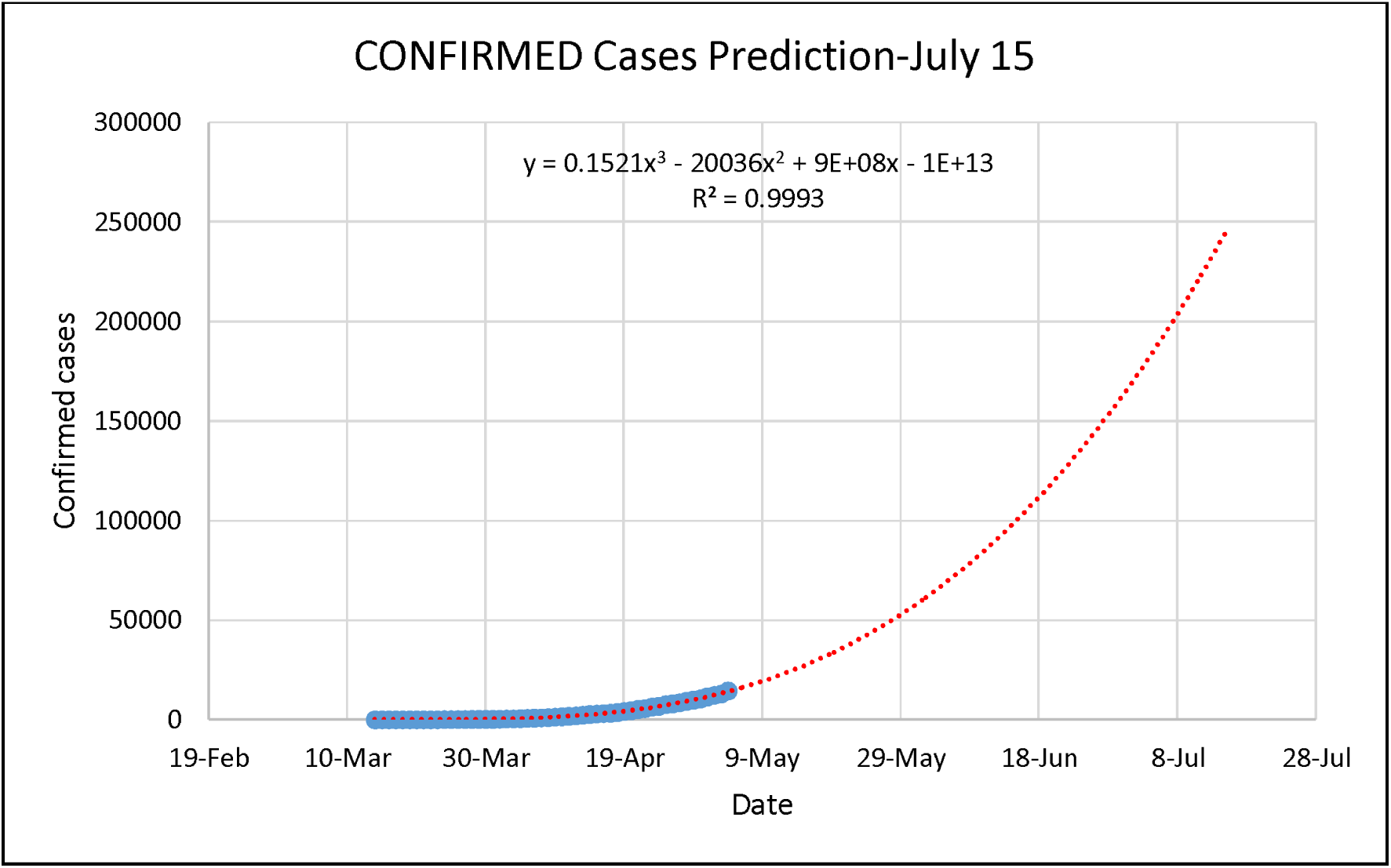
Prediction 15 July.

Continuing with data (14 March 2020-4 May 2020, inclusive of both), hereafter, Mean Square Error and Akaike Information Criterion (AIC) are used as evaluation metrics along with R^2^ for nonlinear regressions due to the highly flexible nature of polynomial curves to fit along with data. Statistics of the model is as shown in table 3.

**Table 3.**
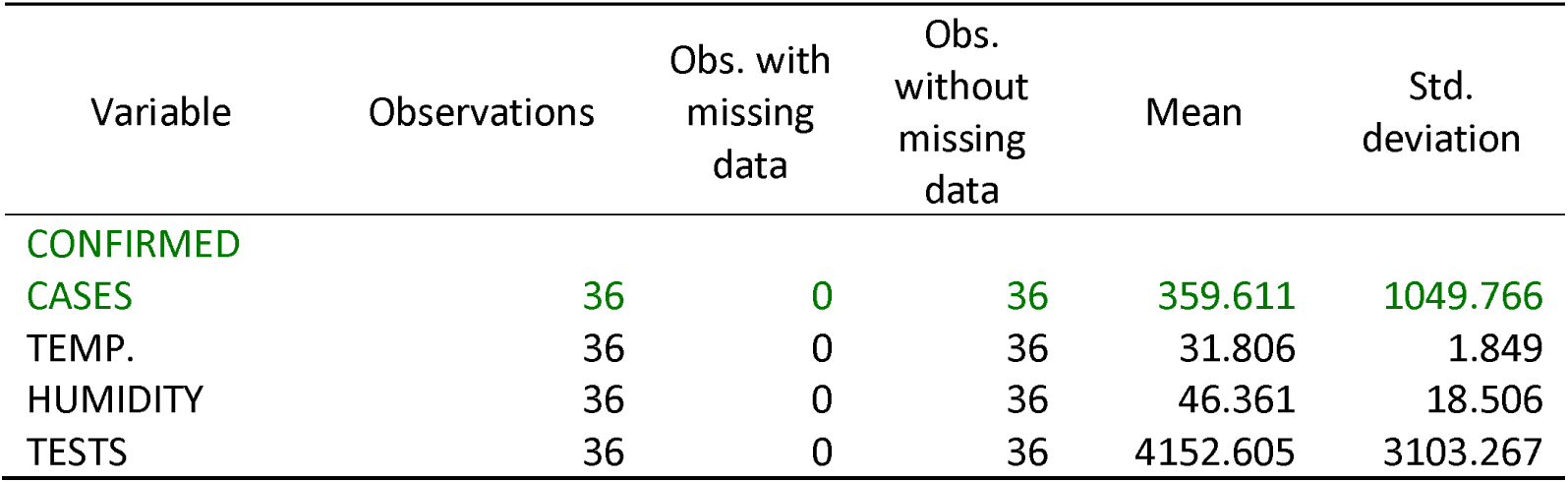
Summary statistics:

| Variable | Observations | Obs. with<br>missing<br>data | Obs.<br>without<br>missing<br>data | Mean | Std.<br>deviation |
| --- | --- | --- | --- | --- | --- |
| CONFIRMED<br>CASES | 36 | 0 | 36 | 359.611 | 1049.766 |
| TEMP. | 36 | 0 | 36 | 31.806 | 1.849 |
| HUMIDITY | 36 | 0 | 36 | 46.361 | 18.506 |
| TESTS | 36 | 0 | 36 | 4152.605 | 3103.267 |

As the observed standard deviations of variables are high, the Grubs tests for the presence of outliers in the observation data are conducted concerning each variable with the two-sided alternate hypothesis and the significance level of 5%.

**Table 4.** Grubbs test for outliers / Two-tailed test (CONFIRMED CASES):

|  |  |
| --- | --- |
| G (Observed value) | 4.525 |
| G (Critical value) | 2.991 |
| p-value (Two-tailed) | < 0.0001 |
| alpha | 0.05 |

Test interpretation:
H0: There is no outlier in the data
Ha: The minimum or maximum value is an outlier
As the computed p-value is lower than the significance level $\alpha=0.05$ , one should reject the null
hypothesis $H_0$ , and accept the alternative hypothesis $H_a$ .

**Table 5.**
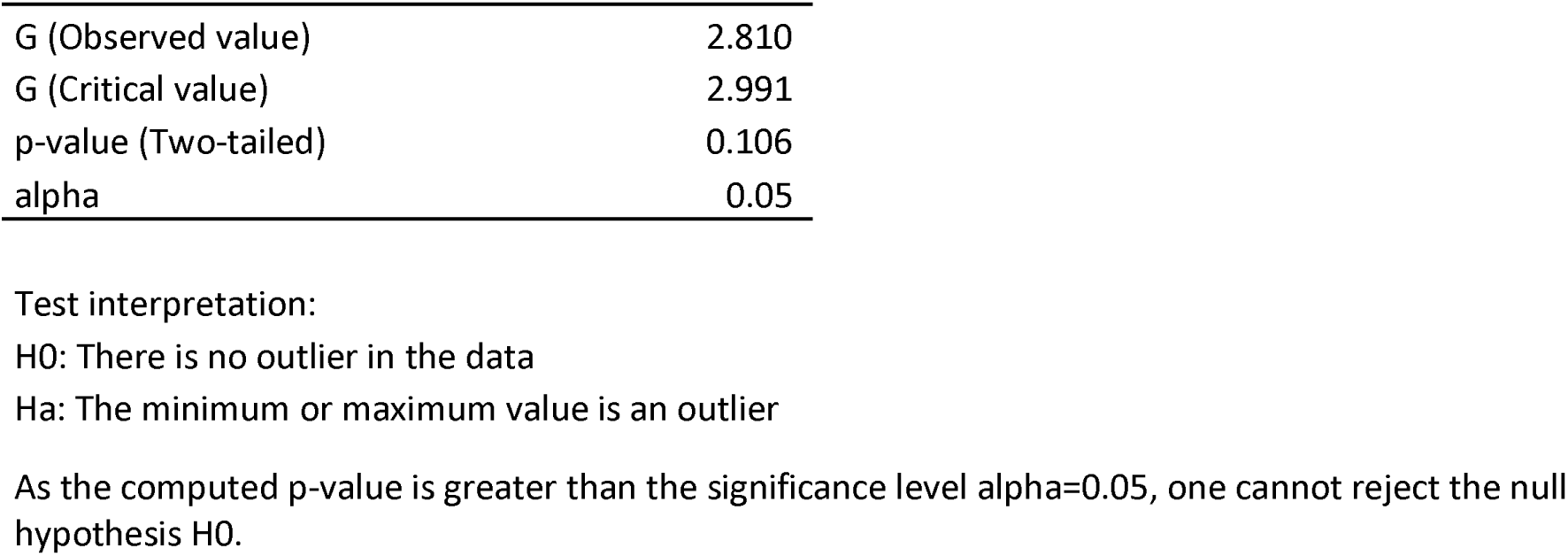
Grubbs test for outliers / Two-tailed test (TEMP.):

**Table 6.** Grubbs test for outliers/ Two-tailed test (HUMIDITY):

|  |  |
| --- | --- |
| G (Observed value) | 1.980 |
| G (Critical value) | 2.991 |
| p-value (Two-tailed) | 1.000 |
| alpha | 0.05 |

Test interpretation:
$H_0$ : There is no outlier in the data
$H_a$ : The minimum or maximum value is an outlier
As the computed p-value is greater than the significance level $\alpha=0.05$ , the null hypothesis $H_0$ cannot be rejected.

**Table 7.** Grubbs test for outliers/ Two-tailed test (TESTS):

|  |  |
| --- | --- |
| G (Observed value) | 3.284 |
| G (Critical value) | 2.991 |
| p-value (Two-tailed) | 0.013 |
| alpha | 0.05 |

Test interpretation:
$H_0$ : There is no outlier in the data
$H_a$ : The minimum or maximum value is an outlier
As the computed p-value is lower than the significance level $\alpha=0.05$ , the null hypothesis $H_0$ is rejected, and the alternative hypothesis $H_a$ is accepted.

As evident from tests conducted for outlier detection, there are highly maximum and highly minimum values present in data of confirmed cases and tests per million. These values occur as real-world phenomena that cannot be dropped from the model. Some of the districts are profoundly affected, and the number of tests per million is high in those respective districts. While few areas have the negligible amount of confirmed cases reported as shown in the Fig. 2. The same is the reason for the higher values of SSE, MSE, and AIC, as shown in Fig.4.

The convergence of points near the origin in the Fig. 10 and some high values of residuals in the Fig. 11 are the effect of outlier observational values due to profoundly affected zones where the number of COVID-19 tests is high. Though these statistically less favorable conditions for a good regression model occur, the presence of outliers is vital as these districts are integral parts of the outbreak region under consideration. Explanatory variables for these outlier observations justify the reason behind the more massive outbreak.

**Fig. 10.**
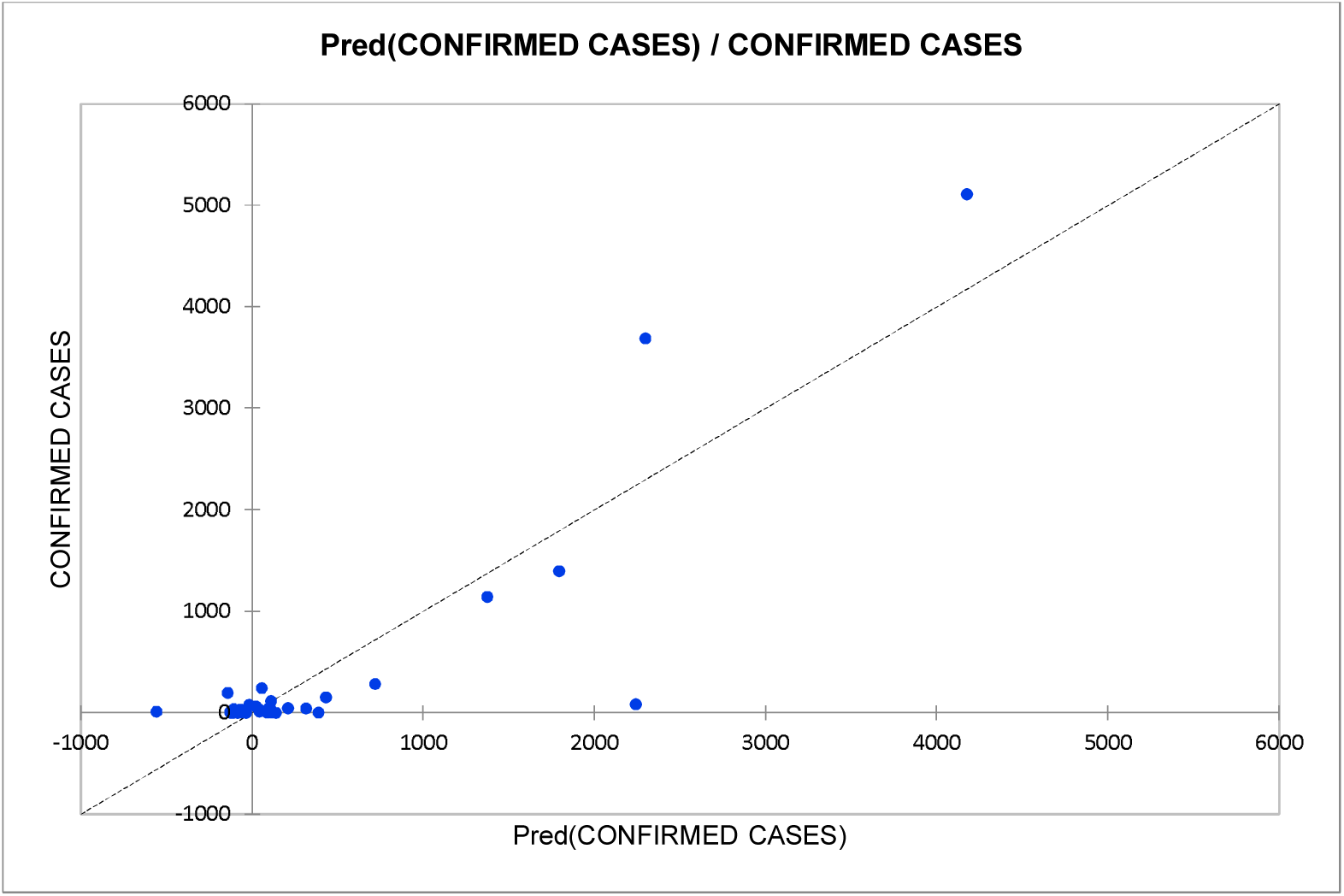
Predicted vs. Actual confirm cases.

**Fig. 11.**
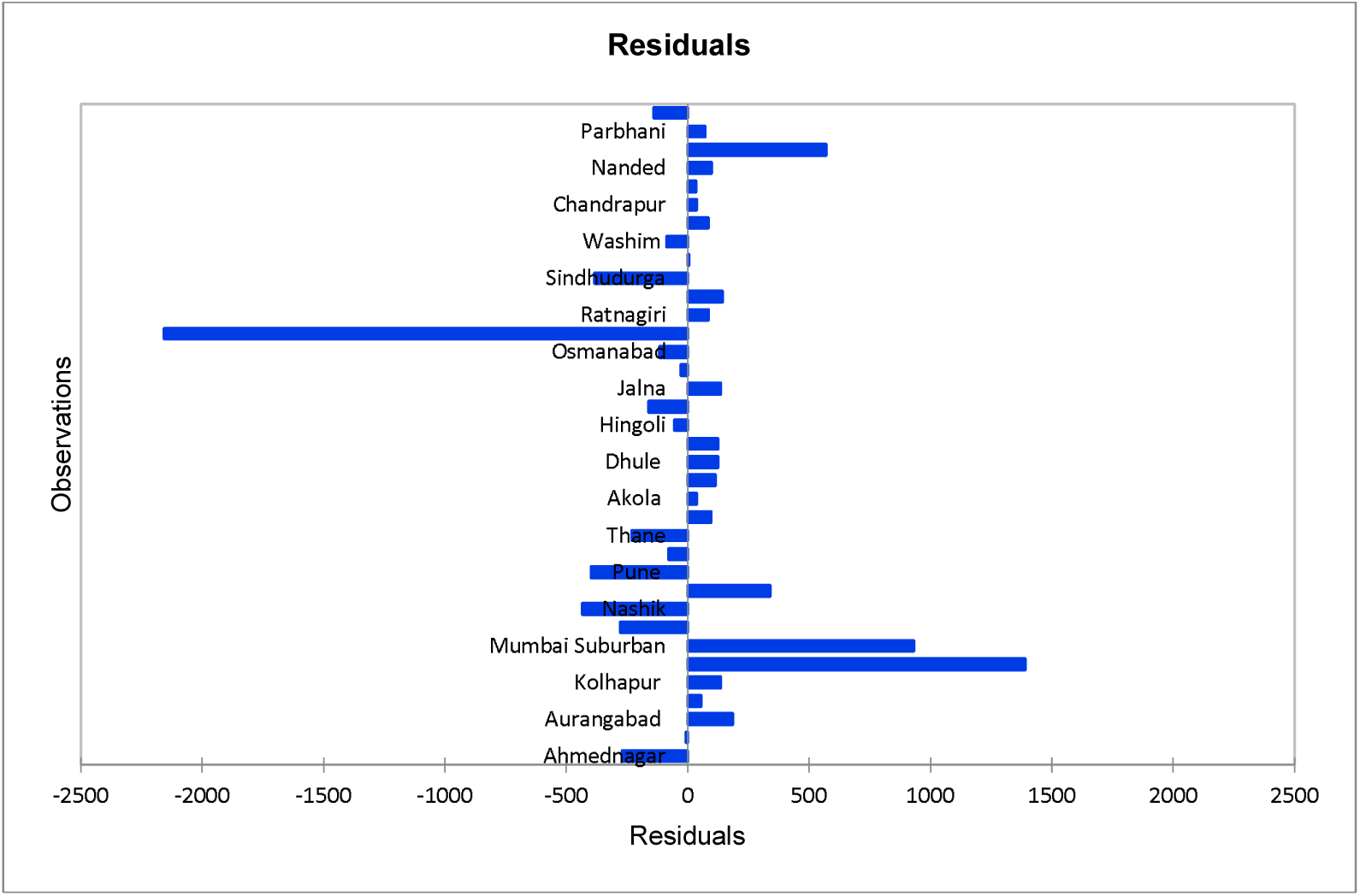
Residuals.

It has been observed that transmission of the virus is significantly high at healthcare-related workplaces such as intensive care units, specialized corona hospitals, quarantine facilities, and housing facilities for medical workers in hotels as well as highly populated public places like government offices, market yards, and transport services. To reduce the rate of transmission in such sites, scrutinizing the impact of explanatory variables on the dependent variable of confirmed cases is crucial. This will lead to estimate possible solutions for the prevention of transmission of the virus under controllable conditions.

The general trend of data in confirmed cases vs humidity plot is observed as an increase in confirmed cases with an increase in humidity in different regions under observation in Maharashtra. This shows a positive correlation between humidity and confirmed cases, as shown in Fig.12. As we traverse from the western coastal area towards the eastern part of the state, a significant drop in humidity is observed with the remarkable lower number of confirmed cases. Whereas in the case of confirmed cases vs temperature, the dependent variable drops significantly low with an increase in temperature of air showing a negative correlation between temperature and confirmed cases as shown in Fig.13

**Fig. 12.**
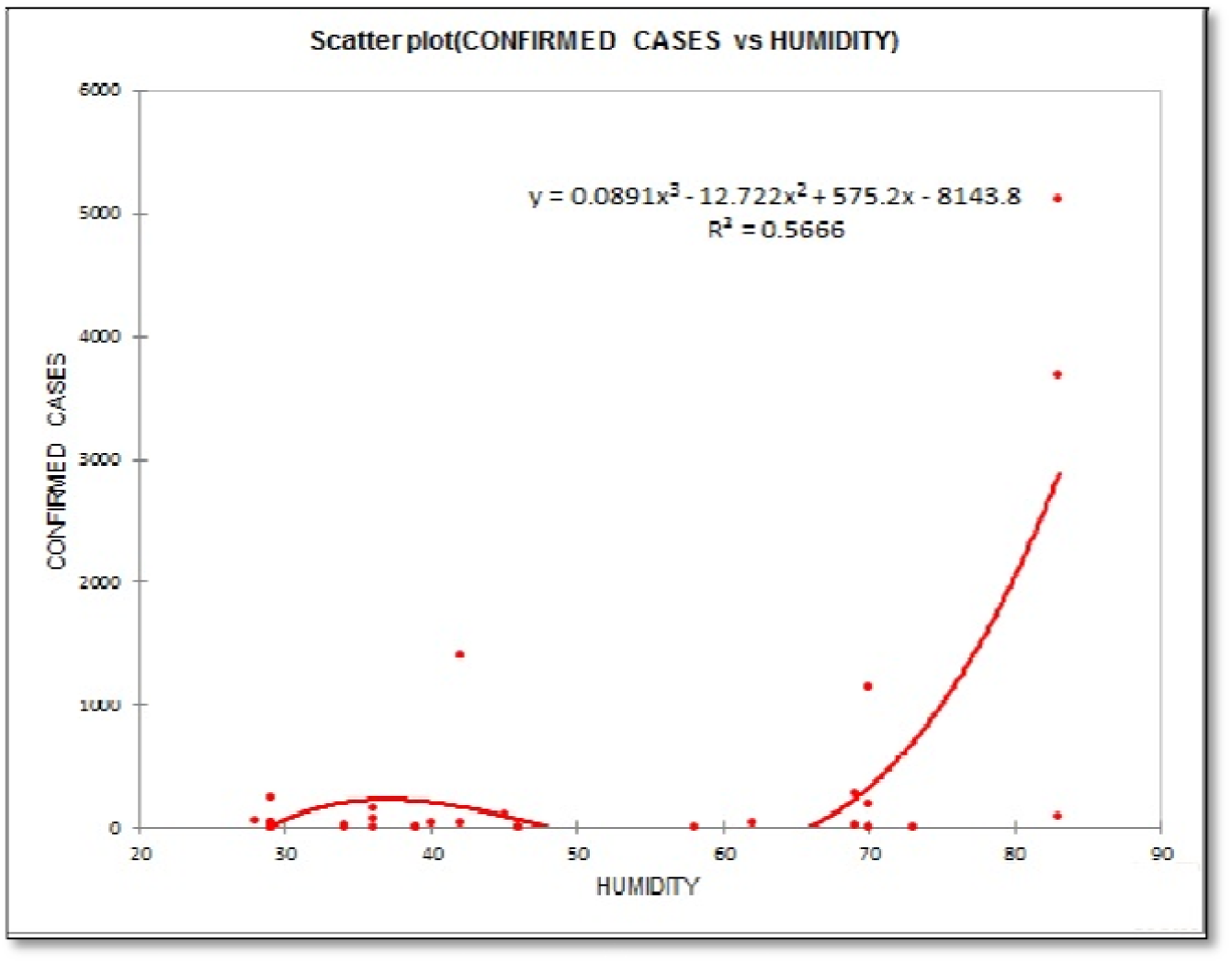
Trend line - confirmed cases vs Humidity.

**Fig. 13.**
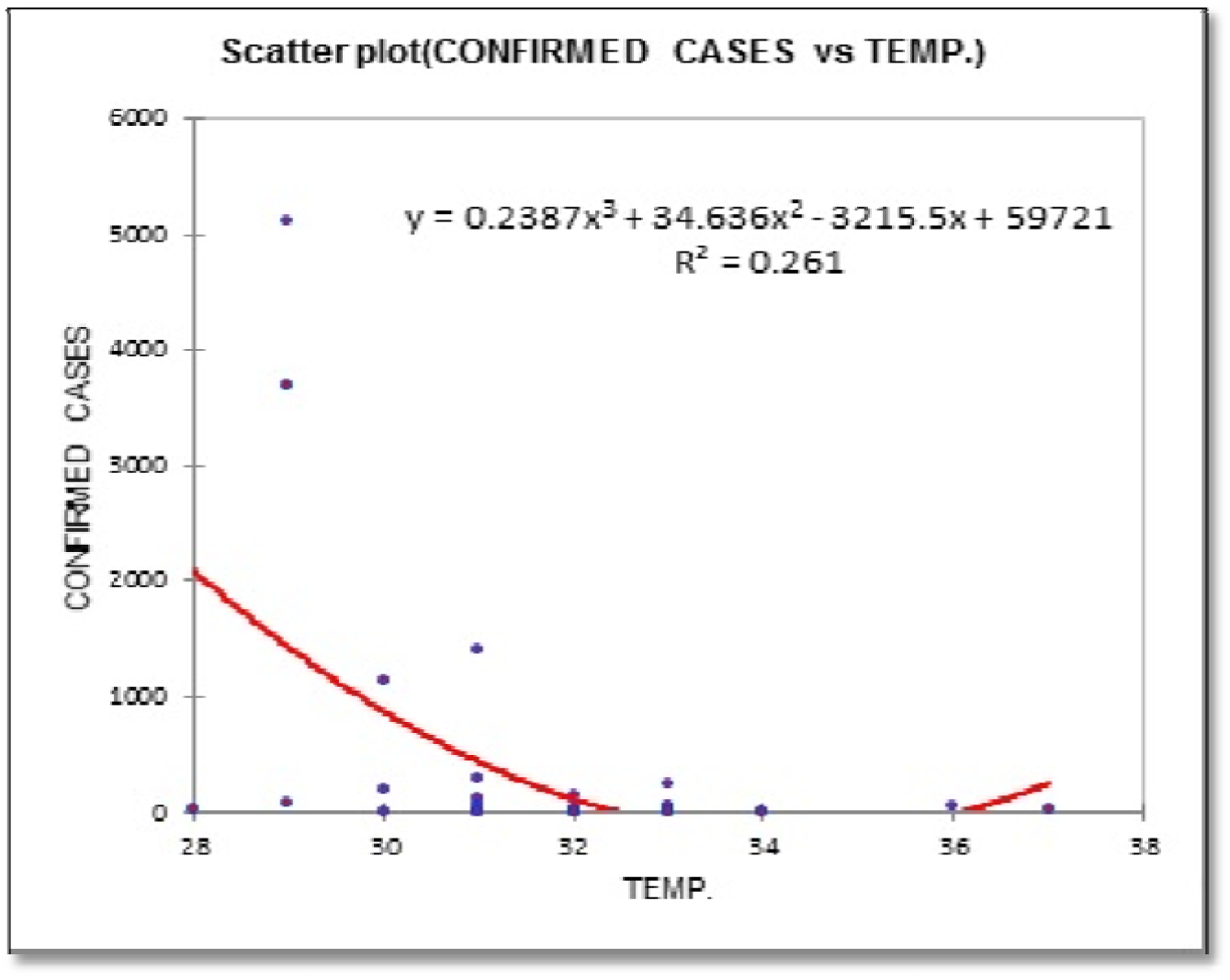
Trend line - confirmed cases vs Temp.

The combined effect of humidity and temperature in the region under consideration through observational data is that relative humidity higher than 60% and temperatures lower than 30°C triggers the spread of the virus. However, higher humidity had proven to be an undesirable condition for the earlier strain of viruses to infect a host. This is contradictory in the context of the present strain of the virus, which is responsible for COVID-19. For earlier strain of the virus, lack of evaporation of aerosol droplets at a lower temperature and higher humidity enabled the settlement of it on surfaces around the infected host. In-activeness of the virus was quick for the earlier case, which is not a case for present virus strain responsible for COVID-19.

Population density is discussed, as shown in the Fig. 14 add the worst effects in the low temperature-high humidity scenario. Proximity living nature in densely populated cities results in a lack of social distancing, which ultimately becomes necessary due to higher humidity enabled settlements of aerosol droplets on surfaces, which are often touched by hundreds of personnel before high humidity prevents droplets from drying out and forming a salt leading to its inactiveness. Thus tests/million, i.e., the number of COVID-19 tests carried out which in turn linearly represents population density triggers the outbreak of this novel coronavirus despite humidity and temperature being in range in which SARS-CoV-1 and SARS-CoV-2 (the different strain of the virus, not same as the present) viruses became inactive to attack a host.

**Fig. 14.**
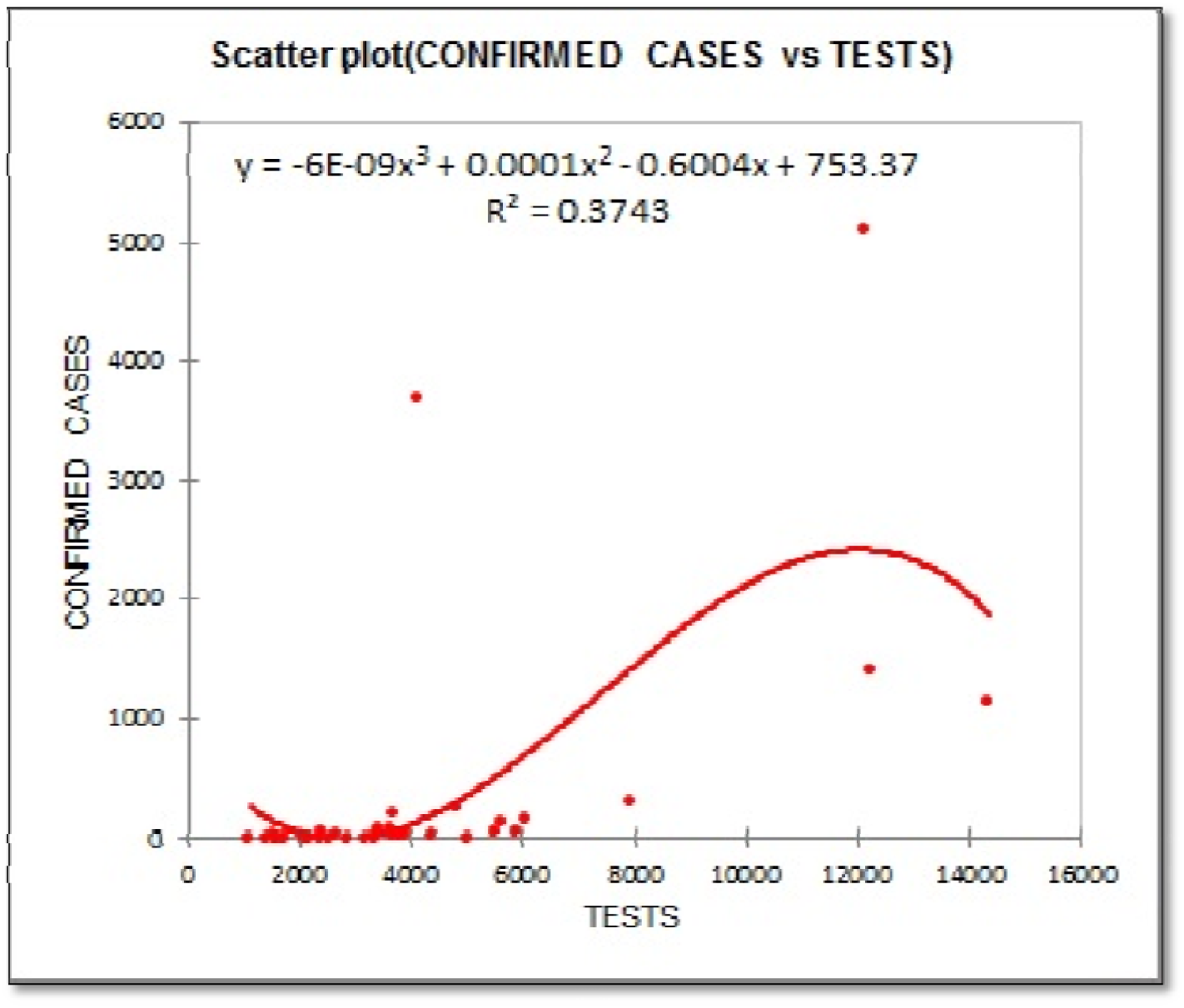
Trend line - confirmed cases vs Tests.

## 4. Conclusion

Third-order polynomial nonlinear regression is evaluated by statistical modelling and validated against real data obtained. Nonlinear regression is carried out to assess the model of an outbreak of novel coronavirus COVID-19 in the state of Maharashtra. Machine learning tool-random forest test is carried out to detect the presence outliers in the data and asses the model as a complex nonlinear model due to retaining outliers, which cause high errors and deviations. Third-order polynomial predicted the outbreak correctly. Based upon this, advanced measures can be taken to have adequate medical facilities in place to withstand a possible outbreak. It is found that despite having unfavorable environment conditions of extremely higher humidity and significantly higher temperatures (for earlier virus strains), an outbreak of COVID-19 is extensive in densely populated areas. This signifies that social distancing, which includes negligible surface contacts, is of utmost importance in public places where humidity and temperatures cannot be controlled. Whereas in specialized COVID-19 dedicated hospitals, clinics, housing facilities, and other indoor areas where surface contacts are significantly more & temperature, humidity can be controlled, these variables must in the precise range which will prevent both; viruses from becoming airborne and also from drying out on the surface and forming salt casing. Humidity less than 60% and temperature higher than 25°C is to be preferred in hospitals to prevent the spread. Dense areas should be vacated, and the corresponding population needs to be relocated to a different region of the state without affecting the concerned natives.

## Data Availability

Data has been taken from the Indian government website for COVID-19 as well as COVID-19 Maharashtra government portal.

https://www.covid19india.org/state/MH

https://arogya.maharashtra.gov.in/1175/Novel--Corona-Virus

## Author Contributions

All author contributions are equal.

## References

1. Dangi, R. R., & George, M. (2020). Temperature, Population and Longitudinal analysis to predict potential spread for COVID-19. Population and Longitudinal Analysis to Predict Potential Spread for COVID-19 (March 24, 2020).

2. Fanelli, D., & Piazza, F. (2020). Analysis and forecast of COVID-19 spreading in China, Italy and France. Chaos, Solitons & Fractals, 134, 109761.

3. Ghazaly, N. M., Abdel-Fattah, M. A., & Abd El-Aziz, A. A. (2020). Novel Coronavirus Forecasting Model using Nonlinear Autoregressive Artificial Neural Network. Journal of advanced science.

4. Ivanov, D. (2020). Predicting the impacts of epidemic outbreaks on global supply chains: A simulation-based analysis on the coronavirus outbreak (COVID-19/SARS-CoV-2) case. Transportation Research Part E: Logistics and Transportation Review, 136, 101922.

5. Luo, J. (2020). When Will COVID-19 End? Data-Driven Prediction. Singapore University of Technology and Design (http://www.sutd.edu.sg).

6. Petropoulos, F., & Makridakis, S. (2020). Forecasting the novel coronavirus COVID-19. PloS one, 15(3), e0231236.

7. Remuzzi, A., & Remuzzi, G. (2020). COVID-19 and Italy: what next?. The Lancet.

8. Guan, W. J., Ni, Z. Y., Hu, Y., Liang, W. H., Ou, C. Q., He, J. X.,… & Du, B. (2020). Clinical characteristics of coronavirus disease 2019 in China. New England journal of medicine, 382(18), 1708–1720.

9. Xu, X., Chen, P., Wang, J., Feng, J., Zhou, H., Li, X.,… & Hao, P. (2020). Evolution of the novel coronavirus from the ongoing Wuhan outbreak and modeling of its spike protein for risk of human transmission. Science China Life Sciences, 63(3), 457–460.

10. Covid19idia.org-Maharashtra “https://www.covid19india.org/state/MH.”

11. Government of Maharashtra Public Health department “https://arogya.maharashtra.gov.in/1175/Novel--Corona-Virus.”

